# Severe Acute Respiratory Coronavirus-2 Antibody and T cell response after a third vaccine dose in hemodialysis patients compared with healthy controls

**DOI:** 10.1101/2022.03.16.22272527

**Authors:** Benedikt Simon, Harald Rubey, Martin Gromann, Astrid Knopf-Völkerer, Boris Hemedi, Sonja Zehetmayer, Bernhard Kirsch

**Affiliations:** Mistelbach-Gänserndorf State Clinic, Institute for Medical-Chemical Laboratory Diagnostics; Mistelbach-Gänserndorf State Clinic, Department for Internal Medicine III – Nephrology and Diabetology; Hainburg State Clinic, Department for Internal Medicine; Center for Medical Statistics, Informatics and Intelligent Systems (Institute of Medical Statistics), MedUni Wien

**Author notes:** Corresponding Author: Benedikt Simon, Liechtensteinstraße 67, 2130 Mistelbach).

**Keywords:** hemodialysis, COVID-19, third vaccine dose

## Abstract

Hemodialysis patients (HD patients) have a high health risk from Severe Acute Respiratory Coronavirus-2 (SARS-CoV-2) infection. In this study, we assess the impact of a third vaccine dose (3D) on antibody levels and T cell response in HD patients and compare the results to those of a healthy control group.

We conducted a prospective cohort study consisting of 60 HD patients and 65 healthy controls. All of them received two doses of the Comirnaty mRNA vaccine and a third mRNA vaccine dose (Spikevax or Comirnaty). The SARS-CoV-2 S antibody response in all participants was measured 6 months after the second vaccine dose and 6 to 8 weeks after administration of the 3D. We also assessed INF-γ secretion 6–8 weeks after the 3D in 24 healthy controls, 17 HD patients with a normal and 20 HD patients with a low or no antibody response after the second dose. The groups were compared using univariate quantile regressions and multiple analyses. The adverse effects of vaccines were assessed via a questionnaire.

After the 3D, the SARS-CoV-2-specific antibody and INF-γ titers of most HD patients were comparable to those of healthy controls. A subgroup of HD patients who had shown a diminished antibody response after the first two vaccine doses developed a significantly lower antibody and INF-γ response compared to responder HD patients and controls, even after the 3D. A new strategy is needed to protect this patient group from severe COVID-19 infection.

## 1. Introduction

Severe Acute Respiratory Coronavirus-2 (SARS-CoV-2) is a coronavirus that has led to a pandemic with global consequences. An infection with this virus can cause severe respiratory failure and death^1^. Patients undergoing hemodialysis on a regular basis (HD Patients) are especially prone to infection with SARS-CoV-2^2,3^ and a severe course of disease with significantly increased mortality^4,5^. Consequently, HD patients are prioritized to receive vaccines against COVID-19. Prior studies^6,7^ showed that HD patients exhibit a diminished antibody response after two vaccine doses. Importantly, a subgroup of HD patients were low/non-responders. The T cell interferon gamma (INF-γ) response in HD patients after vaccination has not been characterized very well, although some studies^8^ aim to improve our knowledge on this topic. In Austria, a third dose (3D) of vaccine has been recommended due to the decline of vaccine-induced antibodies and the rise of COVID-19 cases^9^.

The aim of this study is to measure the impact of the third mRNA vaccine on antibody levels and INF-γ response in HD patients 6 to 8 weeks after the 3D and compare these to healthy controls who also received three vaccine shots. Using such data, we aim to assess the differences between the groups and explore whether further measures are needed to adequately protect this high-risk population.

## 2. Methods

We conducted a prospective cohort study to elucidate the antibody and INF-γ response to vaccination with Comirnaty (BNT-162b2, BioNTech/Pfizer, two doses) and a booster dose of an mRNA vaccine (either Spikevax (mRNA-1273), Moderna or Comirnaty) administered 6 months after the second vaccine dose in HD patients versus healthy controls vaccinated with the same regimen.

### 2.1 Study Population

HD patients were considered eligible if they were on dialysis for at least 3 months and had received vaccination with Comirnaty (vaccination schedule in Section 2.1.3). The healthy control group consisted of volunteer healthcare workers who had been vaccinated using the same regimen. Participants in both groups needed to be 18–99 years old. Pregnant women and individuals with known SARS-CoV-2 infection in the past (diagnosed via patient history and test for nucleocapsid (N) antibody, see Section 2.3.2) were excluded from the study. The study protocol was approved by the local ethics committee. Written informed consent was obtained.

In all, 81 dialysis patients were initially scheduled to receive the 3D. Of these, four contracted SARS-CoV-2 infection, two were not eligible for vaccination due to high CRP values, three received a transplant, and, sadly, 12 passed away (unrelated to COVID-19). Finally, 60 HD patients were included to receive their 3D.

Of these, 21 were identified as low/non-responders (see Section 2.1.1). In this group, two participants (11%) were women and 16 were men. The median age was 72 years (age range 49–82 years).

The other 39 HD patients were designated “responders”. In this group, 15 participants (38%) were women and 24 were men. The median age was 66 years (age range 34 to 83 years). All HD patients received their 3D 6 months after the second dose.

Initially, 80 volunteer healthcare workers were recruited to receive their 3D in the course of the study. Of these, two delayed their 3D and 13 opted not to test for antibodies after the 3D. Finally, 65 healthy controls were included.

This group consisted of 43 (66%) women and 22 men. The median age was 50 years (age range 29–65 years). All demographic data are summarized in Table 1.

**Table 1.** Demographics of the study population. COPD, Chronic obstructive pulmonary disease. RAAS, Renin-Angiotensin-Aldosteron system. CNI, Calcineurin inhibitors. MMF, Mycophenolat-Mofetil. EPO, Erythropoetin.

|  | **Control group (n = 65)** | **HD Patients, responders (n = 39)** | **HD Patients, non-responders (n=21)** |
| --- | --- | --- | --- |
| **Age (y, mean, range)** | 52 (29–65) | 66 (34–83) | 71 (49–82) |
| **Women** | 43 (66%) | 15 (38%) | 2 (11%) |
| **Risk factors** |  |  |  |
| Diabetes | 2 (3%) | 15 (38%) | 8 (38%) |
| COPD | 0 | 10 (26%) | 6 (29%) |
| Hypertension | 18 (28%) | 33 (85%) | 18 (86%) |
| **Primary Kidney Disease** |  |  |  |
| Diabetes | - | 10 (26%) | 7 (33%) |
| Vascular disease | - | 18 (46%) | 3 (14%) |
| Glomerulonephritis | - | 4 (10%) | 4 (19%) |
| unknown | - | 0 | 0 |
| other | - | 7 (18%) | 7 (33%) |
| **Medication** |  |  |  |
| RAAS-Inhibitors usage | 0 | 20 (51%) | 6 (29%) |
| Immunosuppressant usage<br>(Steroids, CNI, MMF) | 2 (3%) | 3 (8%) | 4 (19%) |
| Vitamin D supplements<br>usage | 0 | 28 (72%) | 18 (86%) |
| EPO usage | 0 | 35 (90%) | 18 (86%) |

The medical histories of dialysis patients were extracted from medical records, while the control group was assessed using a standardized questionnaire.

#### 2.1.1 Low/Non-Responders

Twenty-one patients in the HD patient group and none in the healthy control group were identified as low-/non-responders based on their antibody titers being lower than 29 BAU/ml four weeks after the second vaccine, as described in our previous work^7^. Briefly, this cut-off correlated with SARS-CoV-2 neutralization activity of patient sera^10^.

#### 2.1.2 Patient selection for SARS-CoV-2 T cell test

The study participants were assigned an anonymization number at the beginning of the study. Some were then randomly selected to receive an INF-γ release assay (IGRA) test due to the limited availability of test kits. From each of the three subjects’ groups (controls, HD patient responders and HD patient low/non-responders), patients were selected using a random number generator (RANDOM.ORG) set to generate integers that had a maximum value of 80, 60 and 21 (initial numbers of controls, adjusted numbers of HD patient responders and HD patient low-responders, respectively), repeated for the number of IGRA tests available. The participants whose anonymization numbers corresponded to the value generated received an IGRA test 6–8 weeks after their 3D (24 controls, 17 HD patient responders, 20 HD patient low/non-responders).

#### 2.1.3 Vaccination schedules

All study participants were immunized with two doses of Comirnaty timed 3 weeks between the first and second doses. All study participants received 3D 5 to 6 months after the second dose. Of these participants, 43 controls (66%) received Spikevax (Moderna) and 22 controls (33%) received Comirnaty (Pfizer/BioNTech) as their 3D. Of the 39 responder HD patients, 38 received Spikevax (Moderna) and one received Comirnaty (Pfizer/BioNTech). Of the 21 low-responder patients, 15 received Comirnaty and six received Spikevax.

For a graphical representation, see Figure 1.

**Figure 1.**
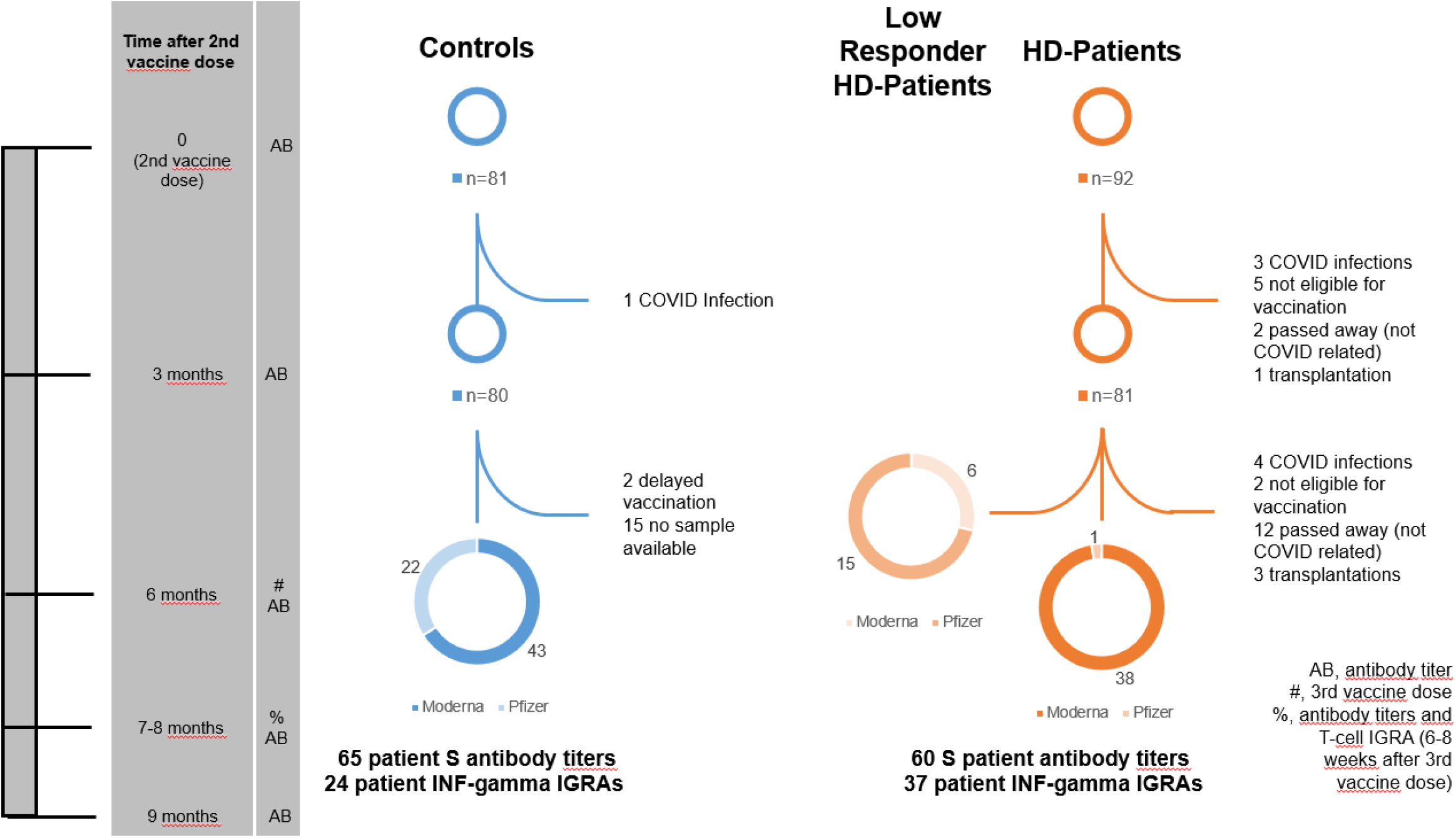
patient flowchart. This flowchart is a graphical representation of the study design. The time axis on the left shows various significant time points in the study for easy orientation. The column beside the time points lists the events that occurred at this time point. Entries in this column are explained in the box at the bottom right of the flowchart.

### 2.2 Antibody titers

#### 2.2.1 Processing of blood samples

Blood draws were performed one week prior to and six to eight weeks after administration of the booster dose. Samples were centrifuged on a Hettich Rotanta 460r centrifuge at 3000 rpm for 10 minutes, aliquoted and anonymized. They were then stored at -70°C and thawed prior to testing.

#### 2.2.2 Serological assessment

All samples were analyzed with an Elecsys®Anti-SARS-CoV-2 test for nucleocapsid (N) antibodies. A positive result in this test led to an exclusion from the study due to a high probability of a past SARS-CoV-2 infection.

The antibody response elicited by vaccination was measured using an Elecsys ® Anti-SARS-CoV-2 S on a Cobas e 801 platform according to specifications, diluted 100-fold. Results were recorded as ranging from 0 (≤0.40 U/ml, lower limit of detection [LOD]) to 25000 (≥25000 U/ml, upper LOD) and assigned to anonymized patient data, as in our previous study^7^. Results showing values greater 15BAU/ml were considered positive.

Longitudinal antibody titers were measured from samples collected 21 days, 3 months and 6 months (last collection on the day of the 3D) after the second vaccine dose. Follow-up samples were taken 6-8 weeks after 3D and 12 weeks after 3D. For the last follow-up, only 16 healthy controls were available.

### 2.3 SARS-CoV-2 S-specific (IGRA)

#### 2.3.1 Processing of blood samples

In the blood draws for antibody titers 6–8 weeks after the 3D, a second sample was drawn from randomly selected patients (see Section 2.1.2) into a lithium-heparin tube. The whole blood samples were incubated in three test tubes lined with SARS-CoV-2 S antigen at 37°C for 24 hours and then centrifuged on a Hettich Rotanta 460r centrifuge for 10 min at 4000 rpm. The supernatant was then analyzed using the ELISA plates provided by the manufacturer.

#### 2.3.2 IGRA test

The T cell response was assessed using the WANTAI SARS-CoV-2 IGRA assay according to specifications ^11^. Twenty µl of specimen diluent and 50 µl of reconstituted standard (400pg/ml in serial dilutions of 1:1, 1:2, 1:4, 1:8, 1:16 and 1:32) and the same amount of serum supernatant was used to fill the wells and incubated at 37°C for 1 h. Fifty µl HRP conjugate was added, and after incubation for 1 hour, the plate was washed 5 times using the washing buffer provided by the manufacturer diluted 1:20 and with 1 min soak time between washes. Chromogen A and B solutions were added to the wells, and after 15 min of incubation in the dark at 37°C, a stop solution was applied to all wells. The plate was read in a dual wavelength photometer (450 and 630 nm). The resulting INF-γ concentrations were recorded as ranging from 0 (≤3 pg/ml, LOD), or any of the conditions specified in the test manual as negative) to 400 (≥400 pg/ml, upper LOD) and assigned to anonymized patient data.

### 2.4 Adverse events

Adverse events (AE) of the 3D for all groups (controls and combined responder and low/non-responder HD patients) were assessed via a standardized questionnaire.

AEs were divided into two categories: local AEs (pain at injection site, redness and/or swelling at injection site and induration at injection site) and systemic AEs (fatigue, headache, muscle and/or joint pain, fever, gastrointestinal symptoms [diarrhea, nausea, vomiting] or other AEs).

Patients were asked to grade their AEs after 3D according to subjective severity. Grading was performed on a scale from 1 to 4. Grade 1 AE signified mild (does not interfere with activity); Grade 2 moderate (interferes with activity); Grade 3 severe (prevents daily activity); and Grade 4 (emergency department visit or hospitalization), analogous to the FDA toxicity grading scale^12^.

### 2.5 Statistics

We investigated the influence of being a member of one of three groups (Controls, HD Patients Low/Non-Responder and HD Patients Responder), sex and age on SARS-CoV-2 RBD-specific antibody titer 6–8 weeks after 3D. Univariate quantile (median) regressions were performed. The quantile regression was chosen due to the skewed distribution of the antibody titer (40 patients with maximum titer observation of 25000BAU/ml, test cut-off). Bootstrap was applied to construct standard errors and perform statistical tests for each independent factor (5000 replications). Pairwise contrasts estimating marginal means (median), standard error (SE) and 95% confidence intervals (CI) were generated for factor variables. P-value adjustment for pairwise comparison of the group variable was performed using the Tukey method. Then, a multiple analysis was computed for all variables with a p-value less than alpha = 0.05 in the univariate analysis.

The influence of sex and age on INF-γ titers was also investigated for the three groups (controls, HD patients responder, HD patients low/non-responder). For this purpose, the same analyses as above were conducted.

The significance level was set to 0.05. The analyses were performed with R 4.1.1 and the R-packages *quantreg* and *emmeans* ^13^.

Boxplots and AE bar graphs were created in Excel 2019 (Microsoft).

### 2.6 Checklist for cohort studies

We used the STROBE cohort checklist when writing our report46.

## 3. Results

### 3.1 Vaccine antibody titer results after three doses

We analyzed the SARS-CoV2 RBD specific antibody titers of 65 healthcare workers and 60 HD patients. Of the control group, 65/65 (100%) had a titer of >15 BAU/ml 6–8 weeks after their 3D. Of the HD Patients, 58/60 (97%) seroconverted after the 3D (3 months after the second dose: 86% HD patients). A graphical representation is depicted in Figure 2 for the healthy controls and in Figure 3 for the HD patients.

**Figure 2.**
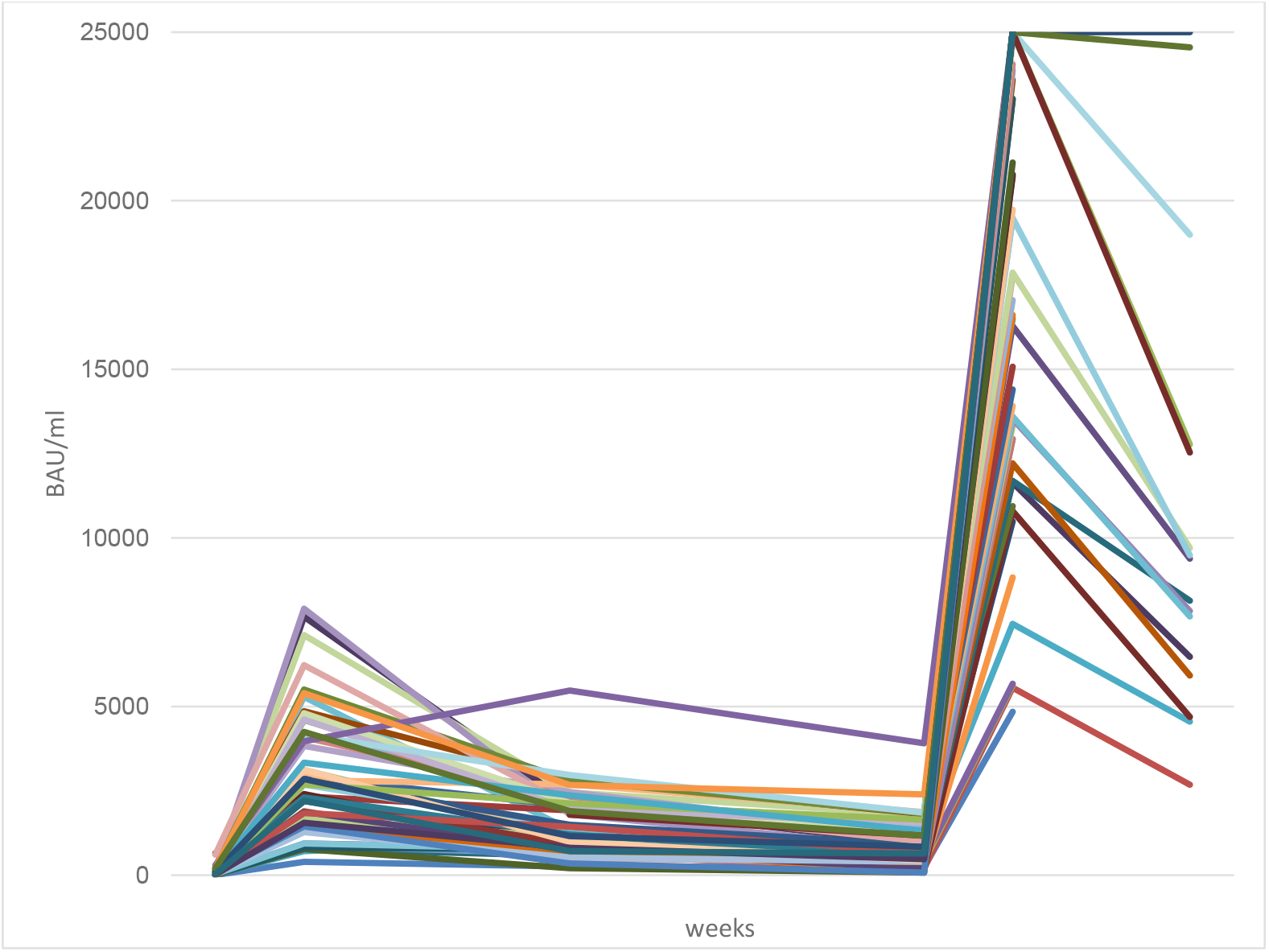
Antibody titers in healthy controls over the course of approximately 1 year. Booster vaccine doses were applied in October 2021. All values in binding antibody units per milliliter.

**Figure 3.**
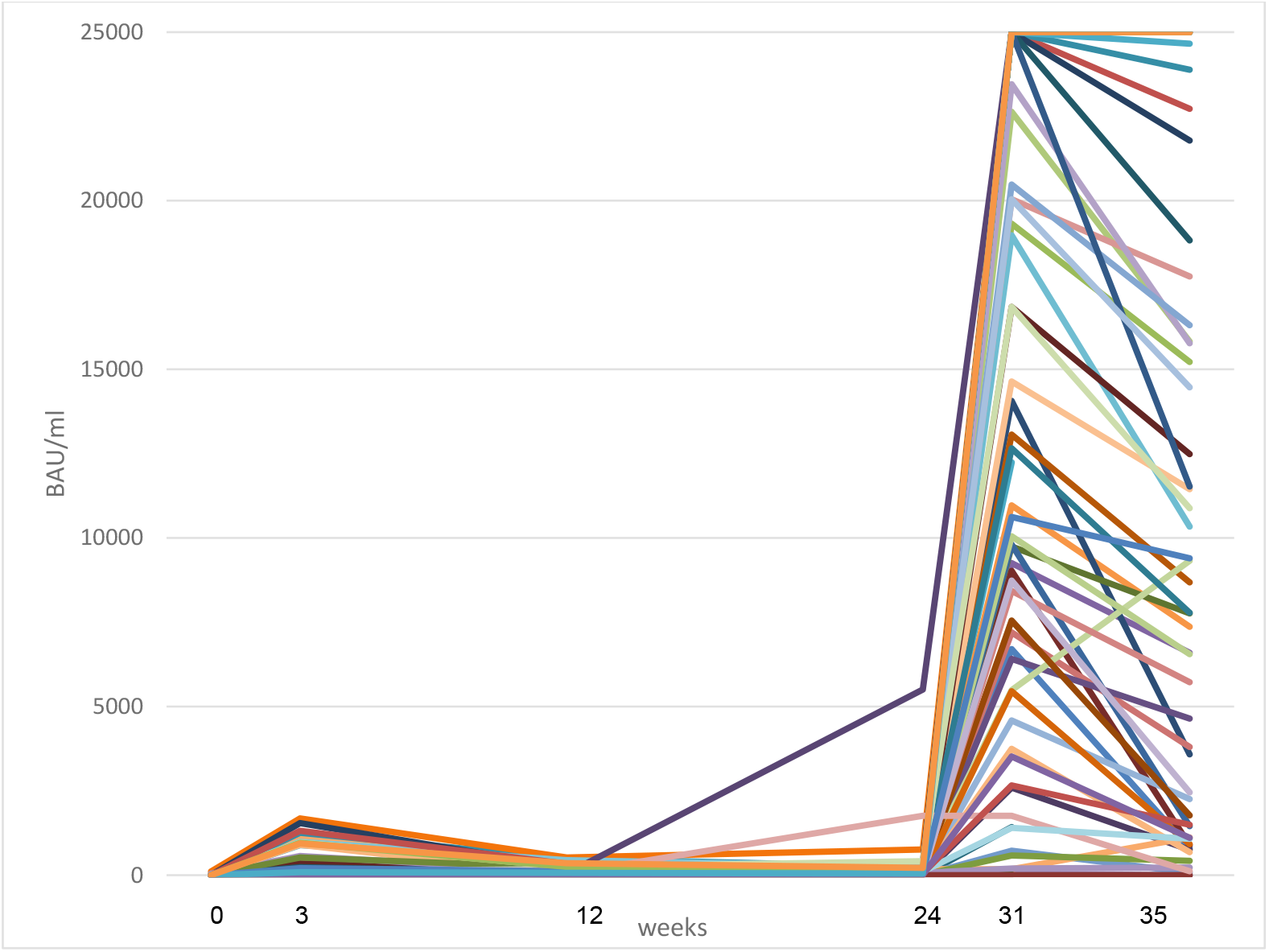
Antibody titers in all hemodialysis (HD) patients (responder and low/non-responder combined) over the course of approximately 1 year. Booster vaccine doses were applied in October 2021. All values in binding antibody units per milliliter.

Importantly, of the 60 HD patients, 21 individuals had an antibody response <29 BAU/ml 3 months after two vaccine doses (“low/non-responders”). Their antibody titer measured 6 weeks after administration of the booster shot showed a certain amount of antibody response. Three HD patients could still be classified as low/non-responders after their 3D. Of these, one suffered from plasmozytoma and another from myasthenia gravis, both conditions with consequences or therapies that could negatively influence B cell function. One low responder achieved an antibody titer of 67 BAU/ml, all 18 remaining patients achieved an antibody titer of 700 BAU/ml or greater (see Table 2), constituting a significant increase caused by the booster vaccine (p < 0.001, Mann-Whitney U).

**Table 2.** SARS-CoV-2 S antibody titers in low/non- responders on the day of and 4 weeks after the booster shot. Patient codes were assigned randomly during anonymization. All titer values in BAU/ml (binding antibody units per milliliter).

| Patient Code | Roche RBD Antibody Titer (BAU/ml) |  |
| --- | --- | --- |
|  | pre-booster | Post-booster |
| 241 | <15 | <15 |
| 209 | <15 | <15 |
| 266 | <15 | <15 |
| 253 | <15 | 67 |
| 262 | <15 | 719 |
| 251 | <15 | 1840 |
| 250 | <15 | 2793 |
| 212 | <15 | 6657 |
| 242 | <15 | 3963 |
| 288 | <15 | 3988 |
| 227 | <15 | 6292 |
| 255 | <15 | 4043 |
| 279 | <15 | 3358 |
| 224 | <15 | 25000 |
| 265 | <15 | 11698 |
| 238 | 19 | 8044 |
| 249 | 23 | 11591 |
| 276 | 24 | 15650 |
| 274 | 26 | 7465 |
| 226 | 28 | 6212 |
| 228 | 28 | 2227 |

When comparing the SARS-CoV-2 RBD-specific antibody response 6–8 weeks after the 3D in the control and HD patient responder groups, no significant difference was observed (p = 0.8), which was in contrast to after the second vaccine dose. However, significantly lower antibody titers were found for HD patients low/non-responders after the 3D compared to the control and HD patient responder group (p < 0.0001, see Figure 4).

**Figure 4.**
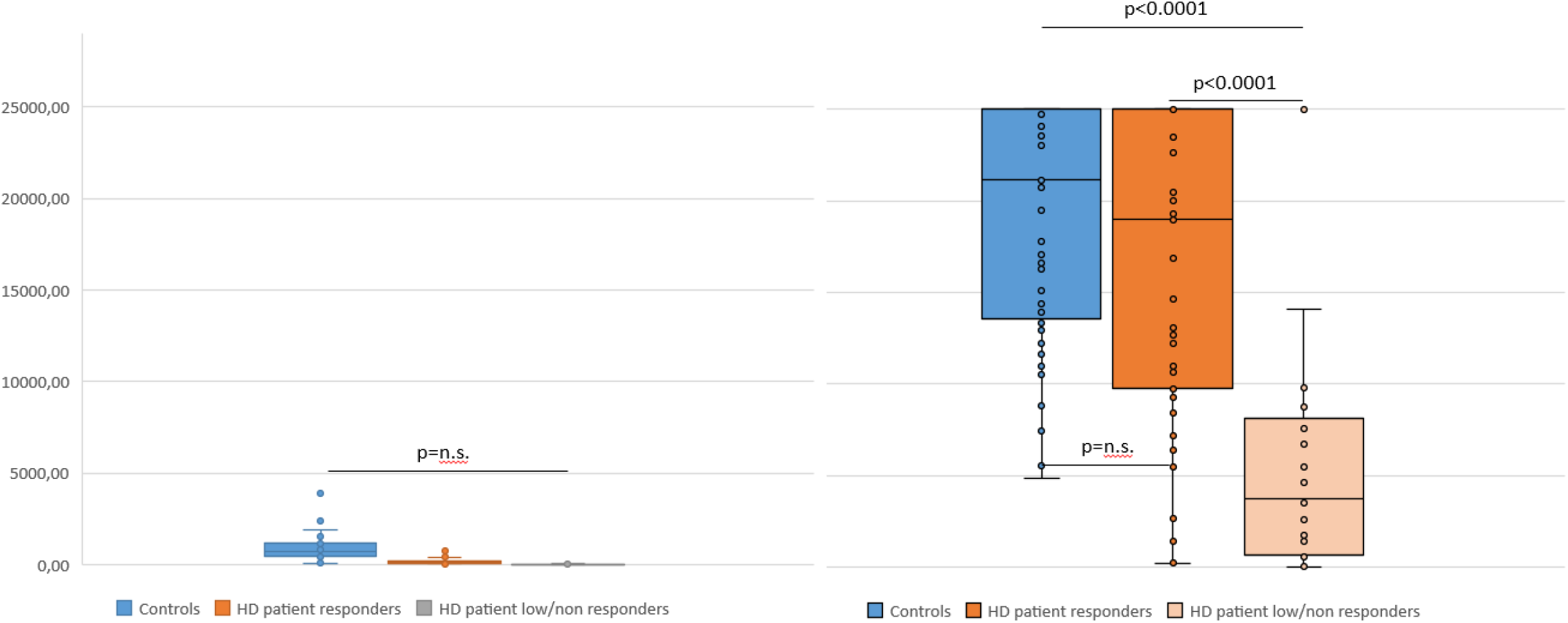
SARS-CoV-2 receptor binding domain-specific antibody titers in Controls and hemodialysis patients (HD Patients, split into responder and low/non-responder groups) 6 months after the second vaccine dose (left) and 6-8 weeks after the third vaccine dose (right). Antibody titres in BAU (binding antibody units) per millilitre. P values <0.05 were considered significant. n.s., not significant. The p value in the left figure is for Controls vs. all HD Patients (responders and low/non-responders combined).

#### 3.1.1 Influences of sex and age on antibody titer

We assessed the influence of variables known to play a role in the antibody response, namely, sex^14^ and age^7,15^ in our groups via a quantile regression test (see Section 2.5). In this study group, men had lower median SARS-CoV-2 RBD-specific titers than women, but this difference was not significant (p = 0.27). Age has a significant influence on antibody titer (p = 0.01): With increasing age, the median antibody titer values decrease. The Spearman correlation coefficient for the two variables age and titer was -0.28.

After including these variables in a multivariate analysis to adjust for sex and age differences in our groups, the differences in antibody titers 6–8 weeks after 3D between the low/non-responders and the other two groups were still significant (p < 0.0001). The control group and the responder HD patients still did not show a significant titer difference (p = 0.43), and age still had a significant influence on the titers (p = 0.33).

**Table 4.** Estimated contrasts of the pairwise group comparison of SARS-CoV-2 antibody titers 6–8 weeks after 3D. All numbers, except for the p-value, are in binding antibody units per ml. P < 0.05 were considered significant. (*), significant value.

| Contrast | Estimate | SE | Statistic | P-value |
| --- | --- | --- | --- | --- |
| Controls vs Dialysis responder | 2158 | 3457 | 0.624 | 0.8071 |
| Controls vs Dialysis low/non-responder | 17386 | 2606 | 6.672 | <0.0001(*) |
| Dialysis responder vs Dialysis low/non-responder | -15228 | 3173 | -4.800 | <0.0001(*) |

### 3.2 INF-γ titers

As a correlate of T cell activation, we compared the SARS-CoV-2-specific INF-γ response in controls (n = 24), HD patient responders (n = 17) and HD patients low/non-responder (n = 20) 6–8 weeks after the booster vaccine. In all, 96% (23/24) of the controls and 76% (28/37) of the HD patients that were tested developed an INF-γ titer greater than the cut-off (3 pg/ml). The boxplots (see Figure 5) show apparent differences in the distribution of INF-γ secretion between the three groups. The quantile regressions results revealed a significant difference only for the median of the control group versus low/non-responder HD patients. However, the sample sizes per group were rather small and the chosen analysis strategy (Tukey test for pairwise comparison) further reduced the power. In the multivariate analysis, neither age nor sex had a significant influence on INF-γ titers.

**Figure 5.**
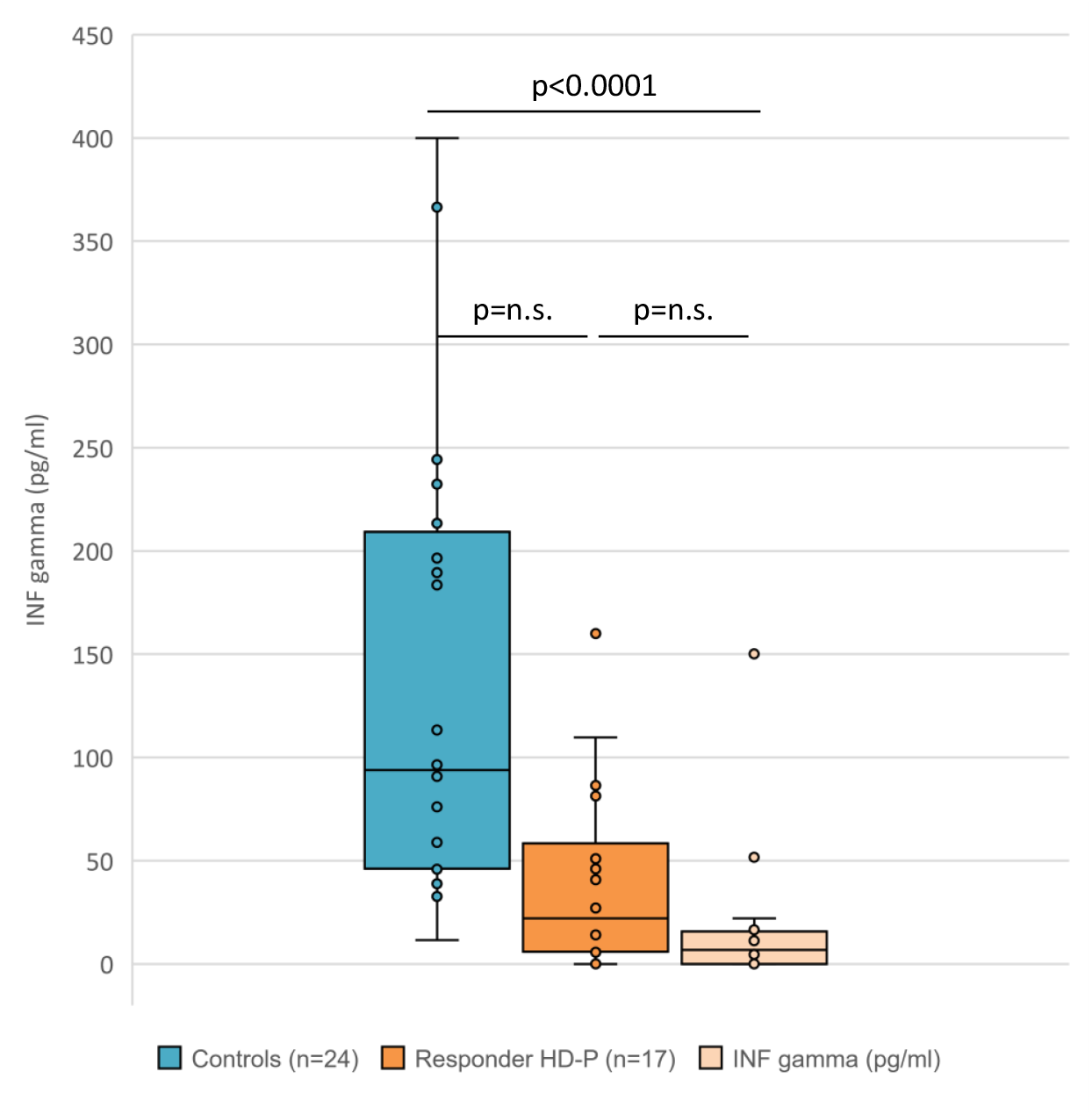
Interferon gamma secretion IGRA titers in Controls and HD Patients (split into responder and low/non-responder) 6–8 weeks after the third vaccine dose. Interferon gamma titers in picograms per ml. HD-P, HD patients. P values < 0.05 were considered significant. n.s., not significant.

### 3.3 Adverse events (AEs) after 3D

Adverse event (AE) reports were analyzed and compared descriptively between the two groups. Sixty-one control group questionnaires and 37 HD patient questionnaires were available. No Grade 4 (emergency department visit or hospitalization) AEs were reported in either group. The control group reported more local AEs after their 3D (67% vs. 62% AEs) and more systemic AEs after the 3D (69% vs 32% AEs) compared to the HD patients (responders and low/non-responders combined) group. See Figures 6a and 6b for a graphical representation.

**Figure 6a.**
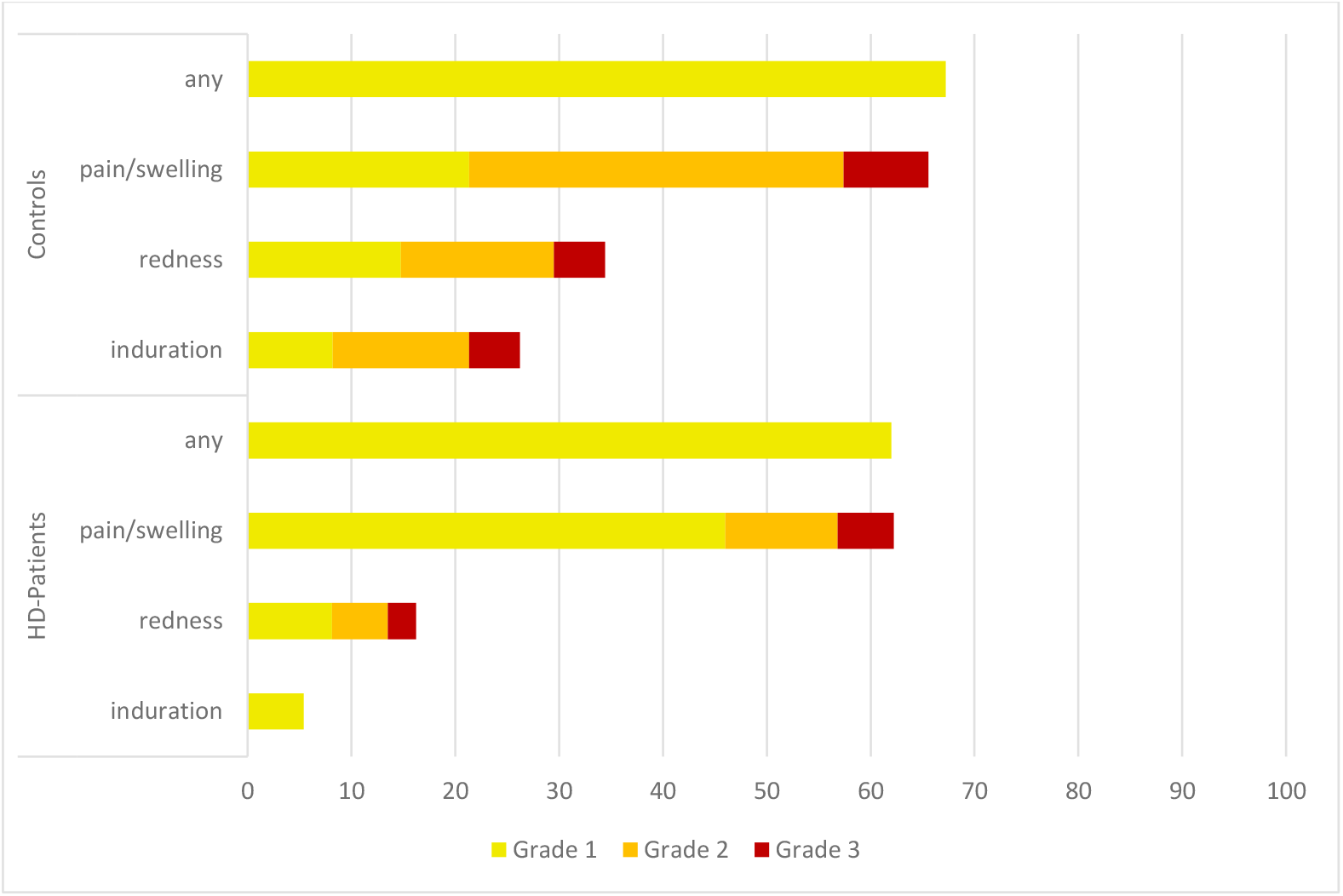
local adverse events (AEs) after the third vaccination. All numbers represent the percentages of dialysis (n = 36) and control (n = 61) patients. The AEs were recorded using a standardized questionnaire and graded by the patients (Grade 1: mild, does not interfere with activity; Grade 2: moderate, interferes with activity; Grade 3: severe, prevents daily activity). No Grade 4 events (emergency department visits or hospitalization) were reported. HD patients, patients on hemodialysis.

**Figure 6b.**
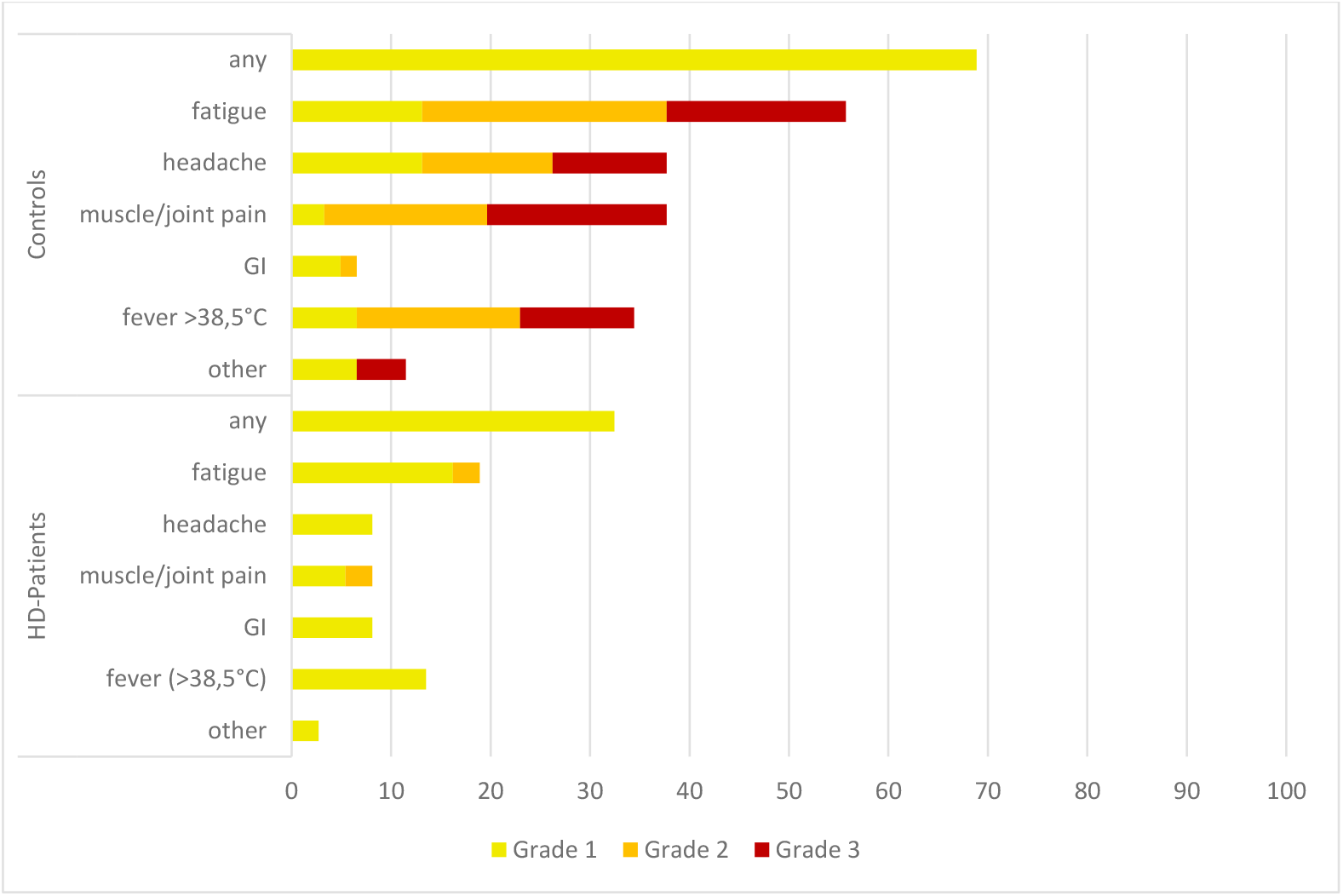
systemic adverse events (AEs) after the third vaccination. All numbers represent the percentages of dialysis (n = 36) and control (n = 61) patients. The AEs were recorded using a standardized questionnaire and graded by the patients (Grade 1: mild, does not interfere with activity; Grade 2: moderate, interferes with activity; Grade 3: severe, prevents daily activity). No Grade 4 events (emergency department visits or hospitalization) were reported. HD patients, patients on hemodialysis; GI, gastrointestinal AEs (diarrhea, nausea and vomiting).

## 4. Discussion

We investigated the SARS-CoV-2-specific RBD titers in healthy controls and HD patients 6–8 weeks after 3D. Recently we showed a significant difference in titers between healthy controls and HD patients after two vaccine doses^7^. More HD patients seroconverted after the 3D (97% compared to 86% after the second dose), and the antibody titers did not differ significantly between controls and responder HD patients. Overall, antibody titers, which are an important factor in protection from a severe infection course, improved significantly after 3D in HD patients, even in low/non-responder HD patients. This result contradicts the finding of ^8^, where antibody titers in high responders did not improve significantly; however, the study’s cut-off for low/non-responders was higher than in our design.

We measured SARS-CoV-2 antibody titers without neutralizing capacity or neutralization titers in our study. This could be viewed as a limitation; however, antibody titers are relevant, as demonstrated by recent studies focusing on the correlates of protection from severe infection. In nonhuman primates, after mRNA vaccine immunization^16^ and in adoptive transfer studies^17^, antibodies were identified as one such correlate. The assay we used in this paper, an RBD-specific S antibody ELISA, not only correlates with neutralization tests, it was concluded that S-specific and/or RBD-specific antibody tests can be used as a correlate of protection from severe infection^16^. This conclusion is further reinforced by data on immunization with the AstraZeneca^18^ and mRNA-1273^19^ vaccines. Another study showed a better correlation of the vaccine efficacy of seven different COVID-19 vaccines with binding (S) antibody titers than with neutralization assays^20^. It also provides data from human monoclonal therapy that proves the protective role of antibodies in COVID-19^20^. The RBD antibody test used in our study also showed good correlation with the WHO International Standard and Reference Panel for anti-SARS-CoV-2 antibody^21,22^, thus providing standardized results.

Cut-offs to quantify antibody titers were determined as in our previous study^7^ according to correlation with a neutralization test^10^. Briefly, a serum antibody titer of 29 U/ml 3 weeks after the second vaccine dose was used as the cut-off to divide responder HD patients and low/non-responder HD patients. We feel that these data support the use of antibody titers as a correlate of protection and the cut-off for our groups.

Another possible limitation of our study is the usage of different vaccines (Comirnaty or Spikevax) for the 3D. This circumstance arose due to patient preference and vaccine availability and could potentially lead to differences in antibody titers. Studies show that while Spikevax boosters induce a slightly higher antibody response^23^, the effectiveness of mRNA vaccines against severe disease caused by the SARS-CoV-2 Omicron variant is comparable^24^.

T cell-mediated immunity and immunological memory against SARS-CoV-2 is deemed more robust and longer lasting than antibody levels after infection^25^ and more consistent against variants of concern, also after vaccination with mRNA vaccines^26-31^. It is known that the T cell response has clinical implications. Studies have shown a protective cross-reactivity between SARS-CoV-2 and endemic human coronaviruses for memory T cells^32^. T cell numbers of SARS-CoV-2-specific T cells in peripheral blood predict protection in individuals with low anti-S IgG responses^33^. IGRA assays can be used as a clinically relevant marker of T cell activation^6^. Vaccines induce a highly conserved T cell immunity that is able to neutralize the Omicron^34,35^ and other variants, potentially even future mutations^36^. In our study, although most of the study participants who were tested (96% of the control and 76% of the HD patients group) developed a SARS-CoV-2-specific INF-γ response after the 3D, INF-γ titers were significantly lower in low/non-responders HD patients. As the Omicron variant rises to become the dominant pandemic strain, this could become a potential problem. SARS-CoV-2 Omicron seems to have developed a significant level of immune evasion against antibodies elicited by other variants or vaccines, effectively reducing the neutralizing capacity of serum^37^, but the virus epitopes targeted by T cells remained relatively constant. Studies show a conserved T cell response against Omicron, whereas B cell/antibody responses against the variant and its sublineages are diminished^38,39^. Thus, T cell responses could be more important in combatting Omicron. Since low/non-responder HD patients’ INF-γ titers are lower than those of controls and responder HD patients, they may be even more susceptible to symptomatic and severe infection and/or have a higher mortality than controls and responder HD patients when confronted with this virus variant.

One limitation of our study is that our T cell test system tested INF-γ production of cells in the blood of patients. The majority of INF-γ is produced by T cells, but B cells and antigen presenting cells can also secrete INF-γ^40^, and no further differentiation of the T cell response is possible with this test. Nevertheless, in concordance with the literature cited above, we feel confident that the IGRA test can be used as an accurate tool for T cell response and that our test systems are valid surrogate markers for a protective immune response achieved by vaccination.

AEs were less pronounced in HD patients than in the control group; no severe AEs (myocarditis, emergency department visits) were reported. The 3D was well tolerated by both controls and HD patients, which agrees with data from recent studies^8^ and the general safety profile of mRNA vaccines^41^.

In summary, data show that HD patients like those in our study are at a high risk of infection with SARS-CoV-2 and a severe course of disease and mortality.^42-45^ Most HD-Patients developed a robust antibody response after their 3D. Seroconversion improved from 86% to 97% in HD patients. The antibody titers induced by 3D in the HD patient responder group were not significantly lower than the antibody response in our healthy control group after the 3D. Low/non-responder HD patients developed a significantly lower antibody response compared to controls and HD patient responders after the 3D. Nonetheless, their antibody titers improved compared to after two vaccine doses. The responder HD patients’ INF-γ production was equal to that of controls, but low/non-responder HD patients have significantly lower INF-γ production than the other two groups. Taken together, these findings underline that booster vaccination is a valid strategy to enhance immune responses in most HD patients to the level of healthy controls. They also point to a vulnerable group that needs a different vaccine regimen and/or preventative measures beyond vaccination (e.g., masks, social distancing and hand hygiene, testing strategies and patient isolation). Further studies on alternative vaccination strategies (e.g., dosing and scheduling) in HD patients should be conducted to protect this group from severe SARS-CoV-2 infection.

## Supporting information

STROBE checklist for cohort studies

## Data Availability

All data produced in the present study are available upon reasonable request to the authors

